# Biomedical engineers density indicator per hospital beds

**DOI:** 10.1101/2024.09.05.24313063

**Authors:** Luis G. Ayala

## Abstract

A biomedical engineers density indicator per hospital beds (***ϱ***BEhb) is presented. It calculates the ratio between the number of biomedical engineers providing professional services within the hospital setting, either as internal staff or mixed staff as reported by the World Health Organization, and the available hospital beds in a health system for a specific year.

This is an alternative proposal that relates the biomedical engineer to their professional intervention object (medical devices), as opposed to the current measure published by the World Health Organization, which is the density of biomedical engineers per 10,000 inhabitants.

Data on the number of biomedical engineers and hospital beds can be obtained from reports of various state, national, and regional health systems over different time periods.

For the methodological development of the indicator, the text ‘Health Indicators’ from PAHO/WHO has been used as a reference.

## Introduction

Biomedical Engineering (BME) is the branch of engineering dedicated to combining engineering principles with medicine to develop, manage, maintain, and repair technologies and devices for diagnosis, treatment, and rehabilitation of human health (medical device life cycle). This profession is present in public and private hospitals, companies focused on the sales and repair of medical devices (MDs), government regulatory agencies, as well as universities and research centers, both public and private, engaged in the development of new technologies and the training of human capital. (World Health Organization, 2017) (PAHO, 1999) (Gismondi, 2010) (Bronzino, 2006)

Indicators are tools that clarify objectives and impacts, based on pre-established goals and verifiable results. They are designed with specific standards to measure and visualize the progress of processes or proposed goals (Arosemena, 2022). Defining an indicator involves establishing a logical and clear process, which is essential for obtaining accurate data and its correct interpretation. It should have a single objective and specific dimension, with a well-defined name to avoid ambiguities and facilitate calculation (CEPAL, 2017).

The structures of Biomedical Engineering Departments (BEDs) have been developed to meet the needs of their respective healthcare institutions. According to the World Health Organization’s 2017 document (WHO), three basic models have been identified:

- Internal Staff: A group of biomedical engineers who are employed by and work within the hospital, performing tasks such as predictive, preventive, and corrective maintenance of medical equipment, equipment management, purchasing, team management, and oversight of external services.
- Mixed Staff: One person or a group of individuals from the healthcare institution manage contracts and services provided by medical equipment manufacturers or other companies.
- External Staff and Multiple Providers: In this model, the healthcare institution completely outsources the functions typically handled by the BED.

This complexity and variation in the service models for healthcare institutions can be understood from the perspective that BME is a relatively new branch of engineering. Professionals trained in this field have not been directly demanded by the market; rather, they have found opportunities and developed solutions to technical and administrative problems based on their training and skills. (World Health Organization, 2017) (Linsenmeier & Saterbak, 2020)

Studies and analyses have been conducted on how biomedical engineering professionals are educated, structured, organized, and operate, both in general and specifically within BEDs (Abu-Faraj, 2008) (Linsenmeier & Saterbak, 2020) (Newell, 2012) (World Health Organization, 2017) (Gismondi, 2010) (Vergara et al., 2022) (Miller et al., 2023). One such document is the 2017 report by the WHO, which provides a series of definitions and studies on BME concerning its education, professional associations, certifications, legislation, innovation and development of MDs, the role of BEs in the regulation, evaluation, and management of MDs, as well as their role in the evolution of healthcare systems.

This branch of engineering and healthcare has become so important that in Mexico, a legal reform is being discussed to include biomedical engineers within the scope of professional health activities. (Ramírez & Sánchez, 2020)

In the works produced by WHO, indicators for biomedical engineers as well as other professions traditionally related to healthcare, such as doctors and nurses, can be found (WHO, 2006) (WHO, 2006) (WHO, 2019). These indicators are related to the number of people living in a country, state, or territory (P). For doctors and nurses, this relationship is direct between the service provider and the person receiving care. However, biomedical engineers do not provide direct care to individuals; their professional intervention is focused on MDs.

Developing a specific indicator that relates the number of biomedical engineers to their professional intervention object allows for comparing BEs within a healthcare system (HS) over time or even comparing the number of BEs across two or more HS. If this performance information (functional performance metrics, FPMs) is analyzed from all components, the indicator will facilitate improved decision-making in the training, hiring, and specialization of BEs working in a hospital and a specific HS.

BME is a branch of engineering with just over fifty years of existence, which has established professional spaces in both public and private sectors (Linsenmeier & Saterbak, 2020). During this time, pertinent questions about the profession have arisen. Some questions that have arisen as concerns of the profession are: How many engineers are needed in the market? Are there currently more or fewer engineers than required to meet the minimum needs of the healthcare sector? How do we measure this? What minimum knowledge should be acquired in technical instruction for maintenance, management, and administration of medical devices (MDs) at the intermediate or university level? What would be an appropriate professional career path for developing these professionals? How should they be educated? How should their performance be evaluated? These are significant questions about the profession that, once it has reached its development stage, has entered a phase of consolidation, as noted by Miller and colleagues in their 2023 article.

Considering the broad range of professional fields and the diverse areas of professional development that biomedical engineers (BEs) may cover, including research or sales and procurement processes, this work focuses on BEs providing professional services within the hospital setting, specifically within the internal staff or mixed staff models of Biomedical Engineering Departments (BEDs) as reported by WHO (World Health Organization, 2017). Regarding the innovation system in BME or the education of BEs, there are already individuals, groups, and organizations addressing these topics, which are also research lines our group is studying; however, they will not be covered in this work.

This document focuses on providing a tool to measure the number of BEs employed in hospital settings, allowing for comparisons between HS over time, both within themselves and against others.

There are clear and useful precedents established by health institutions. Notably, the WHO has published an indicator for the density of biomedical engineers (***ϱ***BE) per 10,000 inhabitants (WHO, 2019). This organization states that the justification for this indicator is that global health systems need biomedical engineering professionals and technicians to design, regulate, acquire, and maintain medical technologies, and they should be considered part of the health workforce. The calculation involves the ratio of the number of certified biomedical engineers and technicians in a specific geographic area and time period to the estimated population in the same period and location (P), with the result multiplied by 10,000. This calculation is described by formula (a).

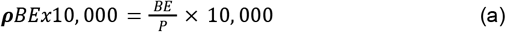

This indicator is based on previous methodologies involving other healthcare professionals who have direct contact with individuals requiring medical care, such as medical doctors and nurses. While BEs are healthcare professionals, they are not directly involved in the diagnosis, care, treatment, or rehabilitation of individuals; their professional focus is on MDs that healthcare professionals use with patients to perform the aforementioned activities. Without these devices, comprehensive care could not be provided.

Therefore, it is necessary to have a tool that correctly relates healthcare professionals to their professional intervention object. It is proposed to modify the measurement approach so that instead of using the human population, MDs are involved in the calculation.

The disparity in professional intervention objects between medicine, nursing, and biomedical engineering (BME) is evident in that 90% of essential actions for universal health coverage can be performed in primary health care units (WHO, 2023). At this level of care, the focus is on prevention, treatment, and management of conditions and diseases that do not require hospitalization. It can thus be inferred that diagnostic, treatment, and rehabilitation medical devices (MDs) are more prevalent in higher levels of care where hospitalization and management of more complex conditions, such as chronic metabolic diseases occur (specialty hospitals).

It is senseless for the reference indicator for BEs to consider only the population and not their professional intervention object. This presents a new problem: with the large number and variety of MDs, how can we account for the number of devices? Is there a reference set of equipment with which we can compare?

A change is proposed in the calculation method for the density of BEs, relating it to hospital beds (***ϱ***BEhb). This indicator is obtained by dividing the number of BEs by the hospital beds (HB) indicator of a HS for a specified time period and geographic location (also known as hospitalization beds or census beds). The formula describing this is expressed in (b):

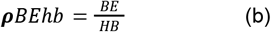

A hospital bed (HB) is defined as a bed installed in hospitals and rehabilitation centers to accommodate patients during observation, diagnosis, care, or treatment. These beds are used to measure indicators such as hospital discharges, occupancy, and length of stay, covering beds for the treatment of both acute and chronic diseases. Hospital beds are considered as the functional units of specialty medical hospitals. For comparison purposes, hospital beds are normalized per 1,000 inhabitants. The indicator of hospital beds per 1,000 inhabitants (HBx1000) is a well-known and continuously updated data point in the health systems of countries and even regions or cities, making the calculation accessible (World Bank, 2011) (National Health System, 1999). The formula describing HBx1000 is:

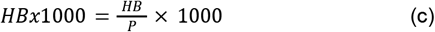

Where HB represents the hospital beds in a HS at a specific time and location, and P is the population at that period. This data is also reported by WHO, but the multiplier factor is 10,000.

The density of biomedical engineers is reported by WHO, and for normalization purposes, a multiplier factor of 10,000 is used. Thus, the available data is the density of biomedical engineers per 10,000 inhabitants (BEx10,000), as expressed in (d). For this indicator, a modification is required: the reported value should be divided by 10 to obtain a measurement comparable with HBx1,000, as shown in (e). This adjustment is not necessary if both indicators originally have the same multiplicative scale.

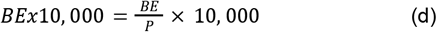

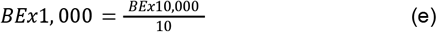

The calculation of the proposed indicator for the density of BEs per hospital beds (***ϱ***BEhb) will require the data on hospital beds per 1,000 inhabitants (HBx1,000) provided by healthcare systems, collected by organizations such as the World Bank or WHO (World Bank, 2011), and the data on BEs per 10,000 inhabitants (BEx10000), which can be obtained from employment data from healthcare systems or from data already compiled by WHO (WHO, 2019). The development of this information is provided in (f).

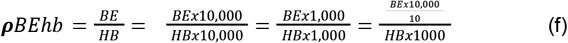

The values for BEs and HBs are originally normalized, and when related, the population (P) becomes a non-relevant data point for calculating the indicator, focusing solely on BEs and their professional intervention object (HBs).

It is relevant to mention that if the HS is robust and includes legislation that considers MDs as essential and necessary for hospital work, and this regulation is adhered to while systematically collecting data on MDs (particularly on HBs and BEs), the indicator will be robust. Conversely, if the HS does not include such provisions or if the collected data lacks rigor, the indicator will present issues with reliability for tracking objectives and decision-making.

## Method

Table 1 presents the technical specifications for the ***ϱ***BEhb developed for this indicator, based on information provided by PAHO/WHO in 2018.

**Table 1.**
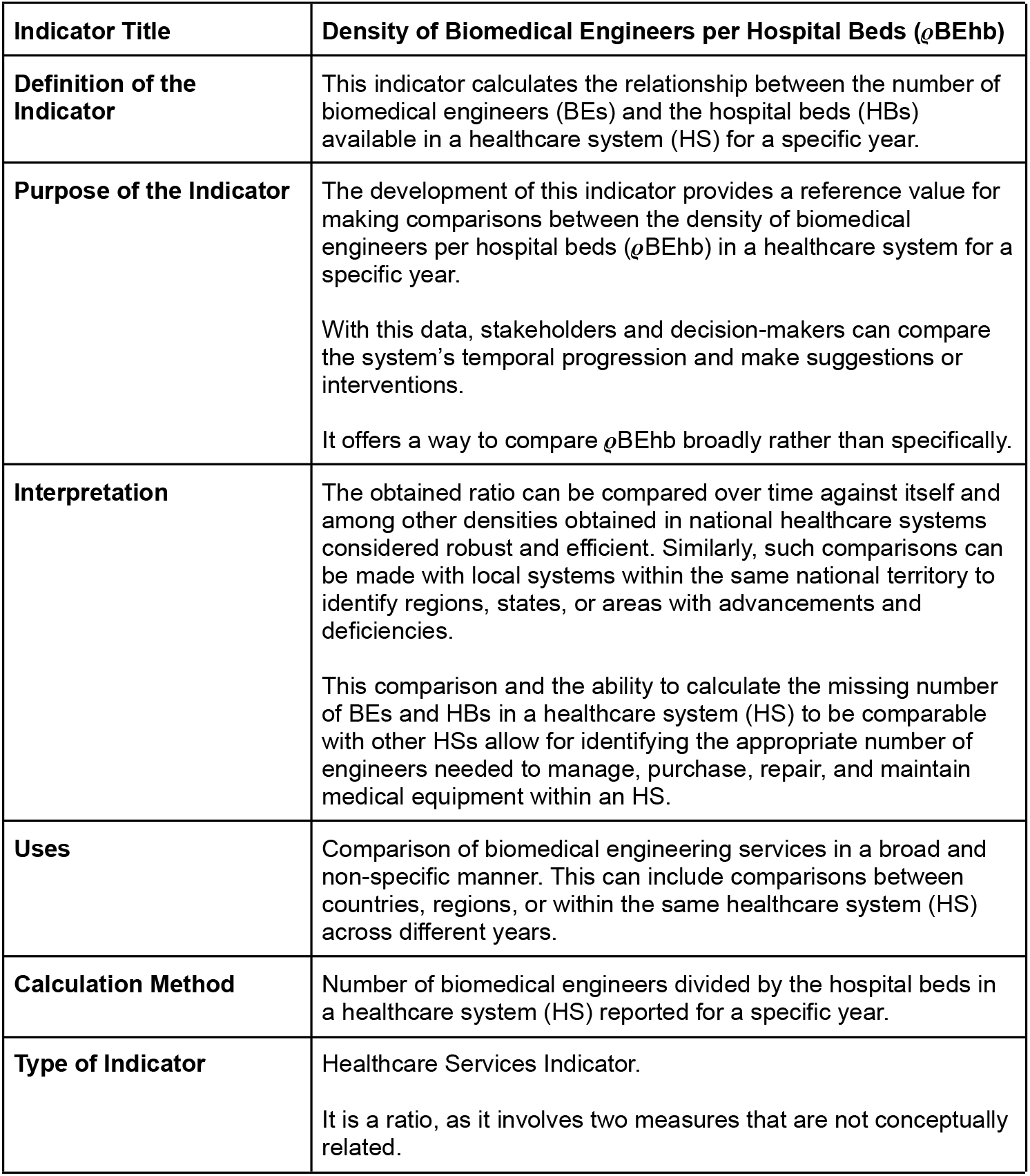
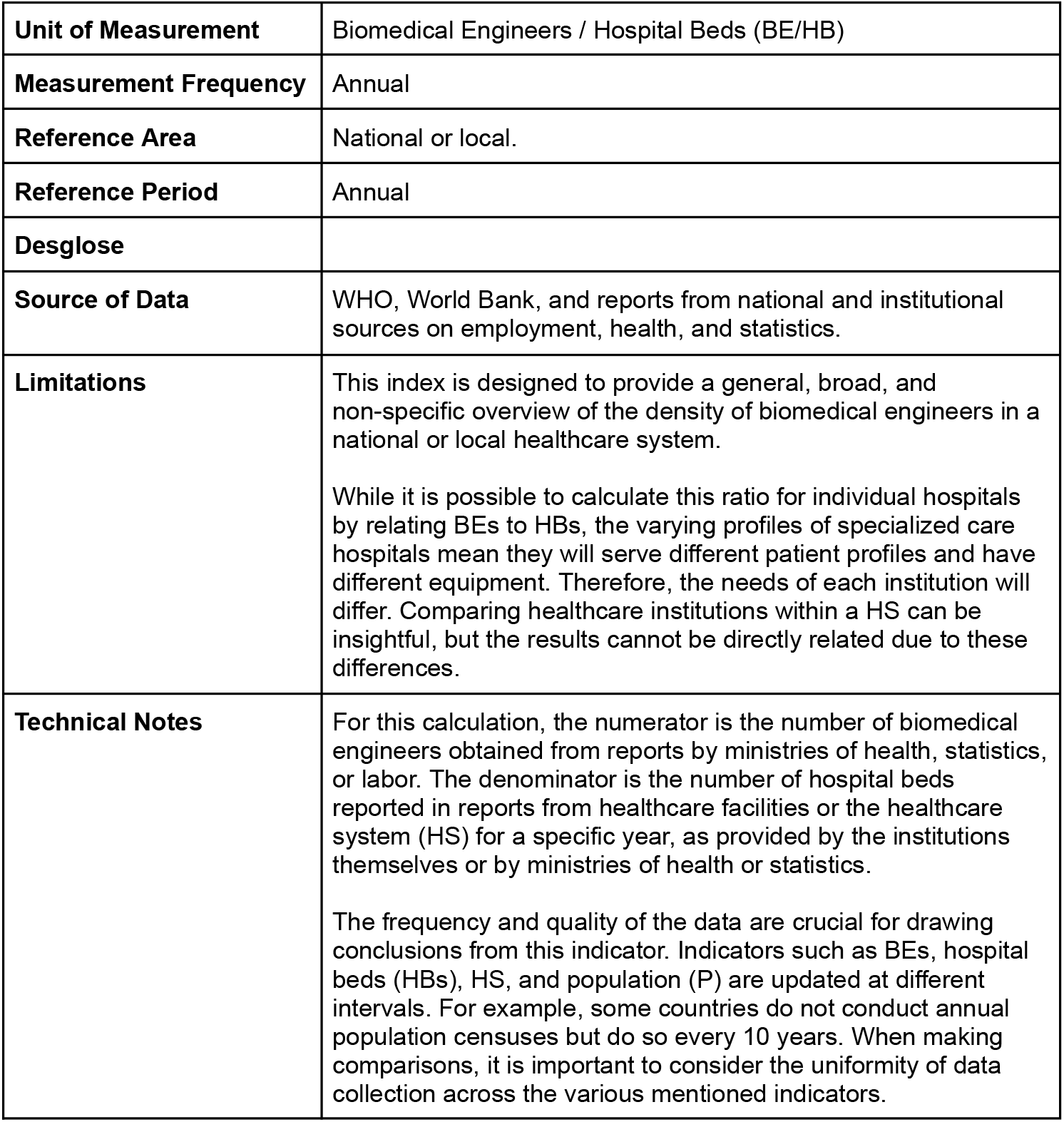
Technical Specification for the Density of Biomedical Engineers per Hospital Beds (***ϱ***BEhb). Source: Authors’ own elaboration.

## Results and Discussion

This work is a proposal that utilizes information from secondary sources for calculating the indicator. Exercises can be conducted based on information published by institutions, health departments, and ministries in countries, regions, and cities around the world.

Table 2 shows the values of ***ϱ***BEhb calculated using (f) based on the available information from the Mexican Ministry of Health and the population (P) data for Mexico (Mx) from the World Bank. This includes the report on existing hospital beds (HB) in the public healthcare system (HS) and the number of biomedical engineers (BEs) employed. To calculate the indicator, it is necessary to obtain the values of BEx1,000 and HBx1,000 using formulas (c) and (e). Notably, the change in ***ϱ***BEhb in the Mexican HS from 2017 to 2022 has generally been positive, with a negative peak of -0.53% from 2017 to 2018 and an increase of 14.41% from 2020 to 2021, which coincides with the onset of the SARS-CoV-2 pandemic.

**Table 2.** Calculation of the Biomedical Engineers per Hospital Bed Density Indicator (***ϱ***BEhb) in Mexico for the Period 2017-2022.

|  | 2017 | 2018 | 2019 | 2020 | 2021 | 2022 |
| --- | --- | --- | --- | --- | --- | --- |
| Population in Mexico | 122,839,258 | 124,013,861 | 125,085,311 | 125,998,302 | 126,705,138 | 127,504,125 |
| Biomedical Engineers (BEs) | 369 | 369 | 415 | 457 | 543 | 601 |
| Biomedical Engineers per 1,000 inhabitants ($\text{BEx}1,000$) | 0.00300 | 0.00298 | 0.00332 | 0.00363 | 0.00429 | 0.00471 |
| Hospital Beds (HB) | 89,085 | 89,562 | 89,538 | 90,555 | 94,047 | 95,780 |
| Hospital Beds per 1,000 ($\text{HBx}1000$) | 0.73 | 0.72 | 0.72 | 0.72 | 0.74 | 0.75 |
| Density of Biomedical Engineers per Hospital Beds ($\rho\text{BEhb}$) | 0.0041 | 0.0041 | 0.0046 | 0.0050 | 0.0058 | 0.0063 |
| Percentage change in $\rho\text{BEhb}$ compared to the previous year | | -0.53% | 12.50% | 8.88% | 14.41% | 8.68% |

Source: Prepared by the authors with data from the Mexican Ministry of Health (2024) and population data from the World Bank (2011).

The values of ***ϱ***BEhb in Mexico for the period 2017-2022 are shown in Figure 1.

**Figure 1.**
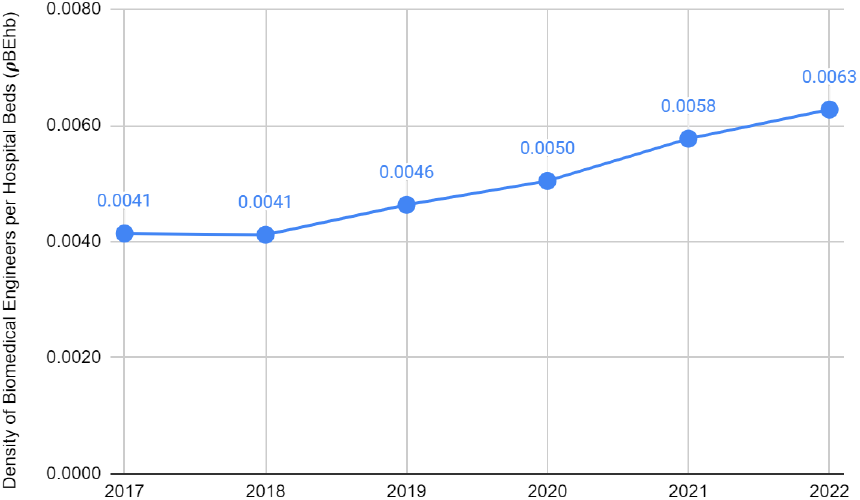
Changes in the Biomedical Engineers per Hospital Bed Density Indicator (***ϱ***BEhb) in Mexico for the period 2017-2022. Source: Prepared by the authors with data from the Mexican Ministry of Health (2024) and population data from the World Bank (2011).

Using data from the WHO for BEs and HB, we have calculated the ***ϱ***BEhb for a selection of countries. The latest reported data for BEs is from 2017, so all data presented will be from that year. Table 2 contains this information and includes the inverse of the ***ϱ***BEhb (***ϱ***BEhb^-1^ = 1/***ϱ***BEhb). The ***ϱ***BEhb can be interpreted as the proportion of BEs per HB; thus, the inverse represents the number of HBs attended to by each BE.

Table 4 contains the biomedical engineer density indicators per 10,000 inhabitants for the same selected countries.

**Table 3.** Calculation of the density of biomedical engineers per hospital beds (***ϱ***BEhb) for a Selection of Countries in 2017.

| Country | $\rho_{BEhb}$ 2017 | $1 / \rho_{BEhb}$ 2017 |
| --- | --- | --- |
| Japan | 0.02475 | 40.4 |
| China | 0.11037 | 9.1 |
| Spain | 0.00741 | 135.0 |
| Denmark | 0.03014 | 33.2 |
| Argentina | 0.00678 | 147.6 |
| Brazil | 0.00218 | 459.4 |
| Costa Rica | 0.01504 | 66.5 |
| USA | 0.02411 | 41.5 |
| Mexico | 0.02422 | 41.3 |

Source: Compiled by the authors with data from (WHO, 2019).

**Table 4.**
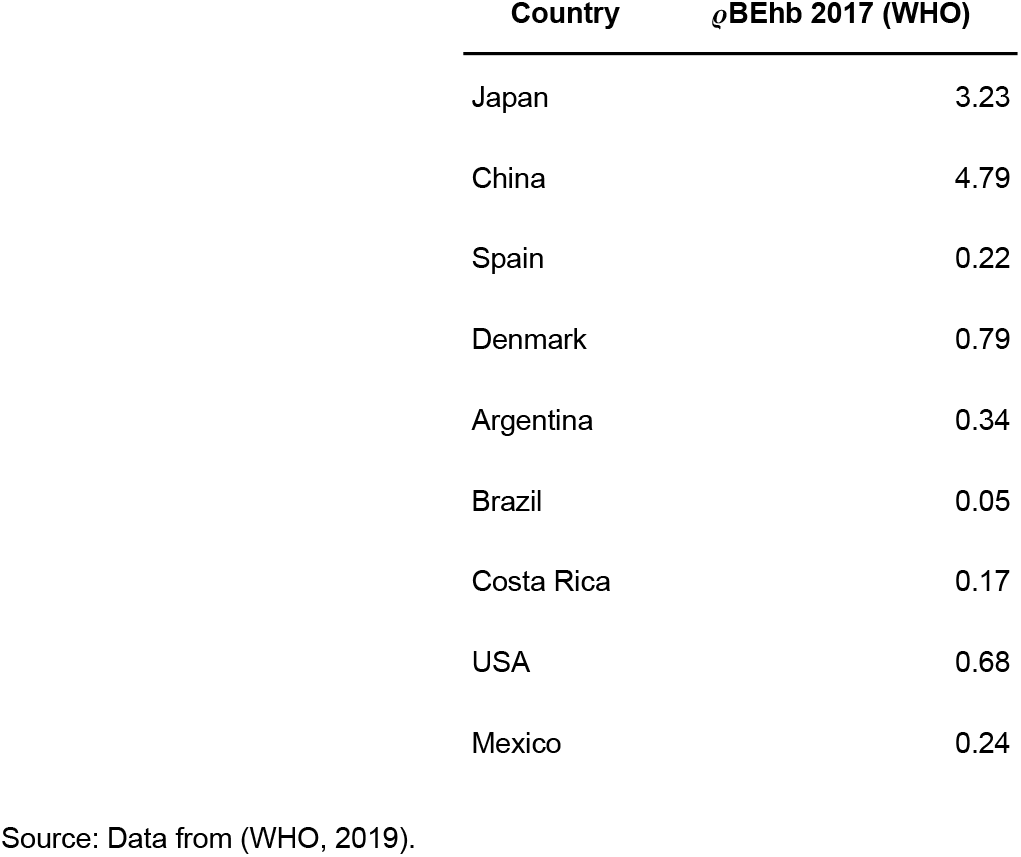
Density of biomedical engineers per 10,000 inhabitants in 2017.

| Country | $\rho_{BEhb}$ 2017 (WHO) |
| --- | --- |
| Japan | 3.23 |
| China | 4.79 |
| Spain | 0.22 |
| Denmark | 0.79 |
| Argentina | 0.34 |
| Brazil | 0.05 |
| Costa Rica | 0.17 |
| USA | 0.68 |
| Mexico | 0.24 |

Source: Data from (WHO, 2019).

When comparing the values shown by both indicators, we can immediately see the difference. The WHO indicator relates BEs to the number of people in a specific location and time, which allows us to estimate how many BEs would serve each person in that population. After normalizing by 10,000, we can determine how many BEs would serve 10,000 individuals. This metric would be relevant if the professional intervention object of BEs were on individuals. Physicians directly care for people, but BEs address the needs of MDs. Therefore, the interpretation of the WHO indicator is less relevant because the demand for BEs is determined by the health system, not the population. While health systems should be designed to serve a specific population size, in practice, this is not always the case, and they operate according to available resources. Some health systems may be under-resourced and unable to meet the required level of care for their population, while others might have excess resources, potentially providing higher quality care.

The proposed indicator (***ϱ***BEhb) allows for a clear interpretation of the results. As shown in Table 3, HS can be compared to determine, for example, that a BE in Mexico (Mx) and in the United States of America (USA) are servicing nearly the same number of hospital beds (Mx=41.3, USA=41.5). Thus, for the size of the health systems in Mx and USA, the number of BEs is comparable. This interpretation cannot be made using the WHO’s indicator of BEs per 10,000 inhabitants, as it only shows a marked difference between Mx (0.24) and USA (0.68), which is dependent on the population size. The WHO indicator might suggest that Denmark (0.79) has a HS most similar to the USA in terms of BEs, but this is not necessarily accurate given the different contexts of the health systems.

In Figure 2 and Figure 3, we visualize the differences between the two indicators.

**Figure 2.**
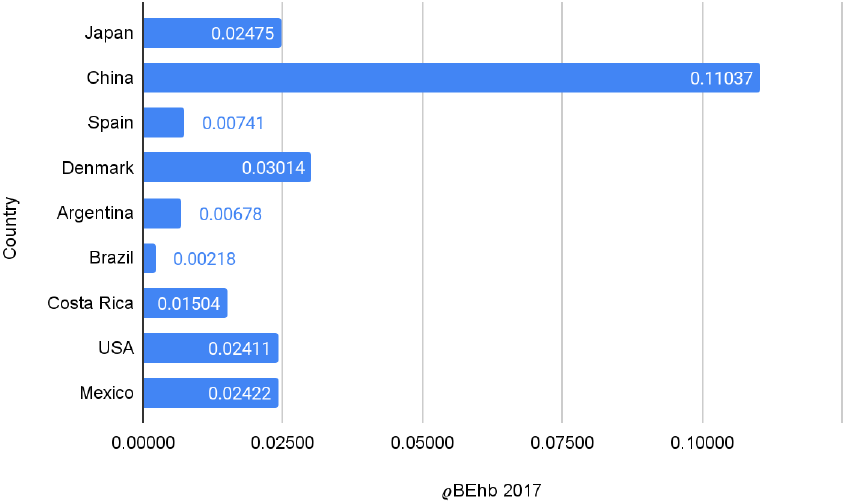
Calculation of density of biomedical engineers per hospital beds (***ϱ***BEhb) in selected countries for 2017. Source: Prepared by the authors with data from (WHO, 2019).

**Figure 3.**
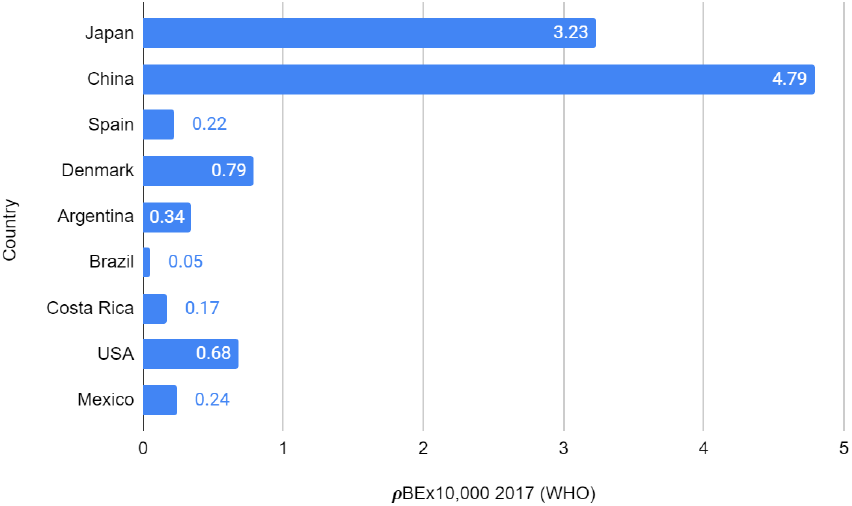
Density of biomedical engineers per 10,000 Inhabitants (***ϱ***BEx10,000) in selected countries for 2017. Source: Prepared by the authors with data from (WHO, 2019).

Another important discussion concerns the adequacy of the number of BEs in a HS. The WHO indicator does not provide insights into this issue. By using the ***ϱ***BEhb indicator, we can assess the relationship between the current HS and a reference HS. For reference, we will use the average number of hospital beds (HB) reported by OECD member countries in their “Health at a Glance 2019” report, as it provides data for the year 2017, which has become our reference year. According to this report, there are 4.7 HBs per 1,000 inhabitants (OECD, 2019). Compared to Mexico’s HB value for the same year, the country would have a deficit of 3.7 HBs per 1,000 inhabitants.

Figure 4 shows the difference in the inverse of ***ϱ***BEhb in Mexico for the year 2017 and compares the value in the healthcare system between the current sector and the reference sector (OECD average).

**Figure 4.**
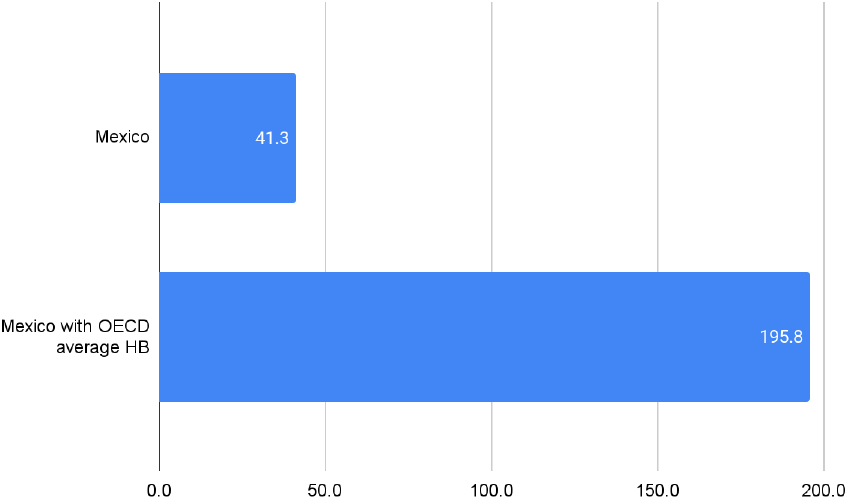
Indicator of biomedical engineers per hospital bed (***ϱ***BEhb) in Mexico with the values of its healthcare system in 2017 compared to the hospital bed value (4.7 per 1,000 inhabitants) in the OECD average for the same year. Source: Prepared by the authors with data from (WHO, 2019) and (OECD, 2019).

If the value of ***ϱ***BEhb were to be maintained (the same proportion of BE to HB), it can be calculated that to handle 4.7 hospital beds per 10,000 inhabitants (the OECD average), Mexico would need to increase its BEs by approximately 11,034 (rounded number). Thus, while Mexico had a comparable number of BE to the USA in 2017, to provide care comparable to the OECD average, Mexico would need to address both a deficit in HB and BE.

The same exercise can be conducted with data from territories and states. Population data can be consulted in censuses, and HB and BE data can be obtained from HS reports. Figure 5 presents the calculation of ***ϱ***BEhb^-1^ for selected states in the USA and Mexico using 2022 data for hospital beds and biomedical engineers, and 2020 data (the closest year) for population.

**Figure 5.**
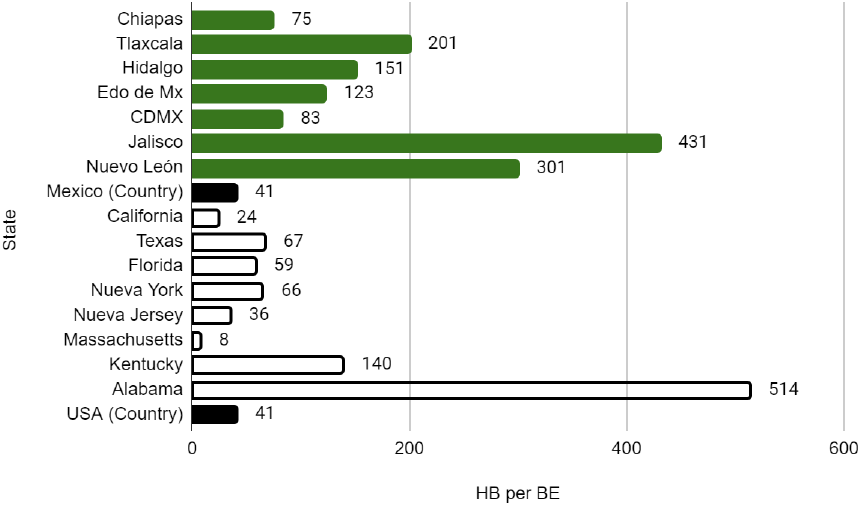
Density of biomedical engineers per hospital bed (***ϱ***BEhb) in selected states of Mexico and the USA for the year 2022. Source: Created by the authors with data from (KFF, 2023), (INEGI, 2020), (United States Census Bureau, 2021), (U.S. Bureau of Labor Statistics, 2023), and (Secretaría de Salud, 2024).

With this state-level information, it is possible to calculate the need for or surplus of biomedical engineers in the state health systems, using the national ***ϱ***BEhb value previously calculated and shown in Table 3 as a reference. Figure 6 displays, in positive numbers, the amount of additional biomedical engineers needed and, in negative numbers, the surplus of engineers required to meet the national value.

**Figure 6.**
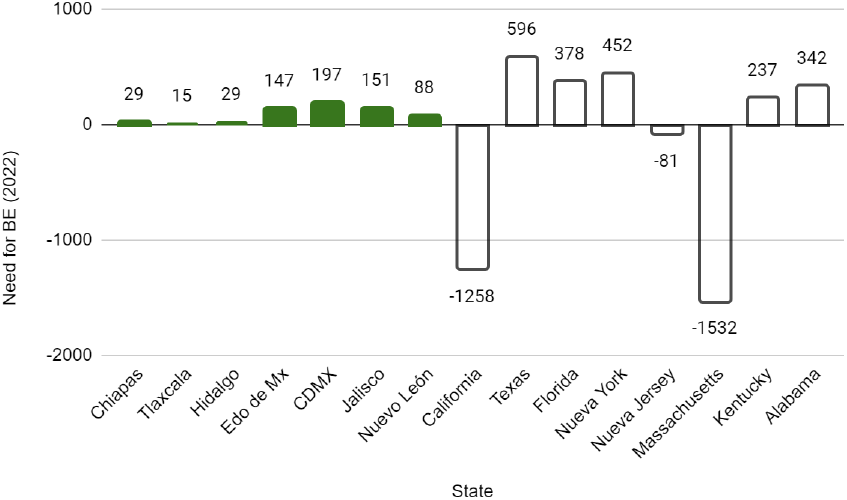
Calculation of the surplus or deficit of biomedical engineers needed in selected states in Mexico and the USA compared to the national ***ϱ***BEhb value. Source: Created by the authors.

Figure 6 shows that, with the exception of California, New Jersey, and Massachusetts, all other state HS in the sampled data exhibit a deficit of BEs. The deficit is more pronounced in the USA state HS compared to those in Mexico, which is a direct consequence of the differences in hospital beds (HB) and overall health system resources.

## Conclusions

An indicator for biomedical engineer density per hospital bed (***ϱ***BEhb) has been developed, based on recommendations from the Pan American Health Organization and the World Health Organization.

Comparisons of healthcare systems over time and against other systems have been effectively conducted. Additionally, a hypothetical scenario was simulated to determine whether a specific healthcare system would have a deficit or surplus of biomedical engineers.

The indicator can be used as a calculation tool for planning the hiring and education of biomedical engineers in national or local healthcare systems.

The value of the indicator depends on the state of the healthcare system being analyzed. A healthcare system might yield ***ϱ***BEhb values comparable to those of systems in other countries recognized for their high-quality healthcare and lead to the conclusion that the number of biomedical engineers is sufficient to meet its needs. However, this does not guarantee that there are no deficits in professionals, infrastructure, or resources within the analyzed healthcare system.

The analysis of ***ϱ***BEhb in the Mexican healthcare system shows that the number of biomedical engineers is comparable to that in the USA. However, when examined from the perspective of how it should perform relative to its population size and international references, the results indicate that Mexico has a deficit in both hospital beds and biomedical engineers.

## Data Availability

All data produced are available online at:
https://data.worldbank.org/
https://www.who.int/data/gho/data/indicators
https://www.inegi.org.mx
https://www.kff.org
https://www.census.gov
https://www.bls.gov
http://www.dgis.salud.gob.mx

## Acknowledgments

I would like to thank the METDI team and the Biomedical Engineering Department at INCMNSZ for their feedback on the ideas in this work. Special thanks to Juan Jesús Mejía Fernández, José Francisco Muñoz del Ángel, Ricardo Bautista Mercado, Nadezhda Aguilar Blas, César Alexis Vega Pelayo, and Lisset Flores Moreno.

## Notes

### Competing Interest Statement

The authors have declared no competing interest.

### Funding Statement

This study did not receive any funding

